# Hand-Crafted Quantitative Radiomic Analysis of Computed Tomography Scans Using Machine and Deep Learning Techniques Accurately Predicts Histological Subtypes of Non-Small Cell Lung Cancer

**DOI:** 10.1101/2024.03.20.24304608

**Authors:** Suhrud Panchawagh

**Affiliations:** Smt. Kashibai Navale Medical College & General Hospital, Pune; ReQuir Statistics Solutions

**Keywords:** Lung Cancer, Computed Tomography, Radiomics, Histopathology, Artificial Intelligence, Classification

## Abstract

**Background:** Non-small cell lung cancer (NSCLC) histological subtypes impact treatment decisions. While pre-surgical histopathological examination is ideal, it’s not always possible. CT radiomic analysis shows promise to predict NSCLC histological subtypes.

**Objective:** To use CT scan radiomic analysis from NSCLC-Radiomics data to predict NSCLC histological subtypes using machine learning and deep learning models.

**Methods:** 422 CT scans from The Cancer Imaging Archive (TCIA) were analyzed. Primary neoplasms were segmented by expert radiologists. Using PyRadiomics, 2446 radiomic features were extracted; post-selection, 179 features remained. Machine learning models like logistic regression, SVM, random forest, XGBoost, LightGBM, and CatBoost were employed, alongside a deep neural network (DNN) model.

**Results:** Random forest demonstrated the highest accuracy at 78% (95% CI: 70%-84%) and AUC-ROC at 94% (95% CI: 90%-96%). LightGBM, XGBoost, and CatBoost had AUC-ROC values of 95%, 93%, and 93% respectively. The DNN’s AUC was 94.4% (95% CI: 94.1% to 94.6%). Logistic regression had the least efficacy. For histological subtype prediction, random forest, boosting models, and DNN were superior.

**Conclusions:** Quantitative radiomic analysis with machine learning can accurately determine NSCLC histological subtypes. Random forest, ensemble models, and DNNs show significant promise for pre-operative NSCLC classification, which can streamline therapy decisions.

## Introduction

Lung cancer remains a leading cause of mortality worldwide, with Non-Small Cell Lung Cancer (NSCLC) including adenocarcinoma, large-cell carcinoma, squamous-cell carcinoma (SCC), and undifferentiated carcinoma constituting the majority of all diagnosed cases.^1,2^ Until now, the treatment approach to NSCLC was similar for different histologic subtypes. Early surgical resection with adjuvant chemoradiotherapy has been the mainstay for early stages. Chemoradiotherapy, usually platin-based, with a secondary agent, usually paclitaxel, has been used for advanced stages.^3^ However, different subtypes of NSCLC are associated with distinct patterns of genomic alterations.^4^ Moreover, evidence from clinical trials demonstrates that tumor histology influences response rates, toxicity and progression free survival of targeted chemotherapeutic drugs.^5^ Therefore, histology is now considered an important factor in targeted treatment selection.^6,7^

Traditionally, the gold standard technique for NSCLC subtype identification involves an invasive biopsy procedure, done by a trained pulmonologist or surgeon. This is followed by meticulous histopathological analysis by an experienced pathologist. While invaluable, these methods are time-intensive, carry procedural risks, and are not always conclusive, leaving a gap in time-sensitive and safe diagnostics.^8^ Traditional non-invasive tools to diagnose lung cancer mainly revolve around the detection of biochemical markers.^9^ Radiologic imaging techniques such as computed tomography (CT) has been used as a preliminary or even alternative diagnostic tools for lung cancers, which is currently based on manual reporting.^10^ This leaves a possibility for false negatives in during reporting, which can prove to be costly.^11^ However, CT scans contain rich data that is not apparent to the human eye, but can only be unravelled by studying signal intensities and other such characteristics. This is where the field of radiomics comes into play. Radiomic data extraction refers to the extraction of quantitative features from medical images such as CT scans, essentially converting digital images into minable, high-dimensional data, which offer unique undetected information that can enhance our understanding of the disease and thus provide clinical decision support.^12^ These radiomic features capture normal tissue as well as lesion characteristics, mainly heterogeneity and shape and may be used for clinical problem solving diagnosis by itself or in combination with demographic, histologic, genomic, or proteomic data.^13^ Therefore, in a clinical setting where diagnosticians are busy and time is often of the essence, relying on automated tools like radiomic analysis can help in reducing human time and error and can thus ease bottlenecks in the diagnostic pipeline.

The integration of artificial intelligence such as machine learning/deep learning with radiomics presents an opportunity to harness these vast amounts of data and boost the era of precision medicine. The appeal of a radiomic approach in the context of NSCLC lies not just in its potential accuracy and precision, but also in its non-invasiveness.^14^ By obviating the need for invasive biopsy procedures, patient morbidity can be significantly reduced. Furthermore, by capitalizing on the quantitative nature of radiomics, we can pave the way for more standardized, reproducible, and objective diagnostic criteria that aren’t as susceptible to interobserver variability, which is a challenge with current methods.^15^ The potential ripple effect on clinical practice will be profound, where, for example, initial CT scans done to confirm the presence of a lung nodule could concurrently predict the NSCLC subtype, thereby accelerating the diagnostic journey and ensuring timely and tailored treatment.

With these challenges and potential to overcome them in mind, the intersection of quantitative radiomic features extracted from seemingly unassuming CT scans with artificial intelligence techniques such as machine and deep learning has tremendous potential. Specifically, we focussed our efforts to investigate the efficacy of a hand-crafted quantitative radiomic analysis in predicting the histological subtypes of NSCLC using CT scans. In a landscape where every advancement could mean a significant difference in survival and quality of life, we believe that this approach could make a dent in existing knowledge and clinical practice.

The aim of our study was to evaluate the efficacy and accuracy of hand-crafted quantitative radiomic analysis combined with machine and deep learning techniques in predicting histological subtypes of NSCLC using CT scans. The objectives were to extract quantitative radiomic features from segmented CT scans of patients diagnosed with NSCLC, to categorize and profile these features based on their potential relevance to distinct NSCLC histological subtypes, and thus subsequently train machine and deep learning models that can integrate and interpret the extracted radiomic features. We also validated the performance of these models using a subset of the data, ensuring their predictive accuracy and reliability.

## Methodology

### Study Design

We followed the CheckList for EvaluAtion of Radiomics research (CLEAR) and the Image Biomarker Standardization Initiative (IBSI) guidelines while reporting the results to ensure standardization.^16,17^ Ethical details and eligibility criteria are available from the original study by Aerts et al.^18^ CT scans of 422 patients with NSCLC were used to extract radiomic data. The study was conducted as a retrospective analysis of previously acquired data. A detailed flowchart depicting the technical pipeline from data collection to analysis is provided in Figure 1.

**Figure 1:**
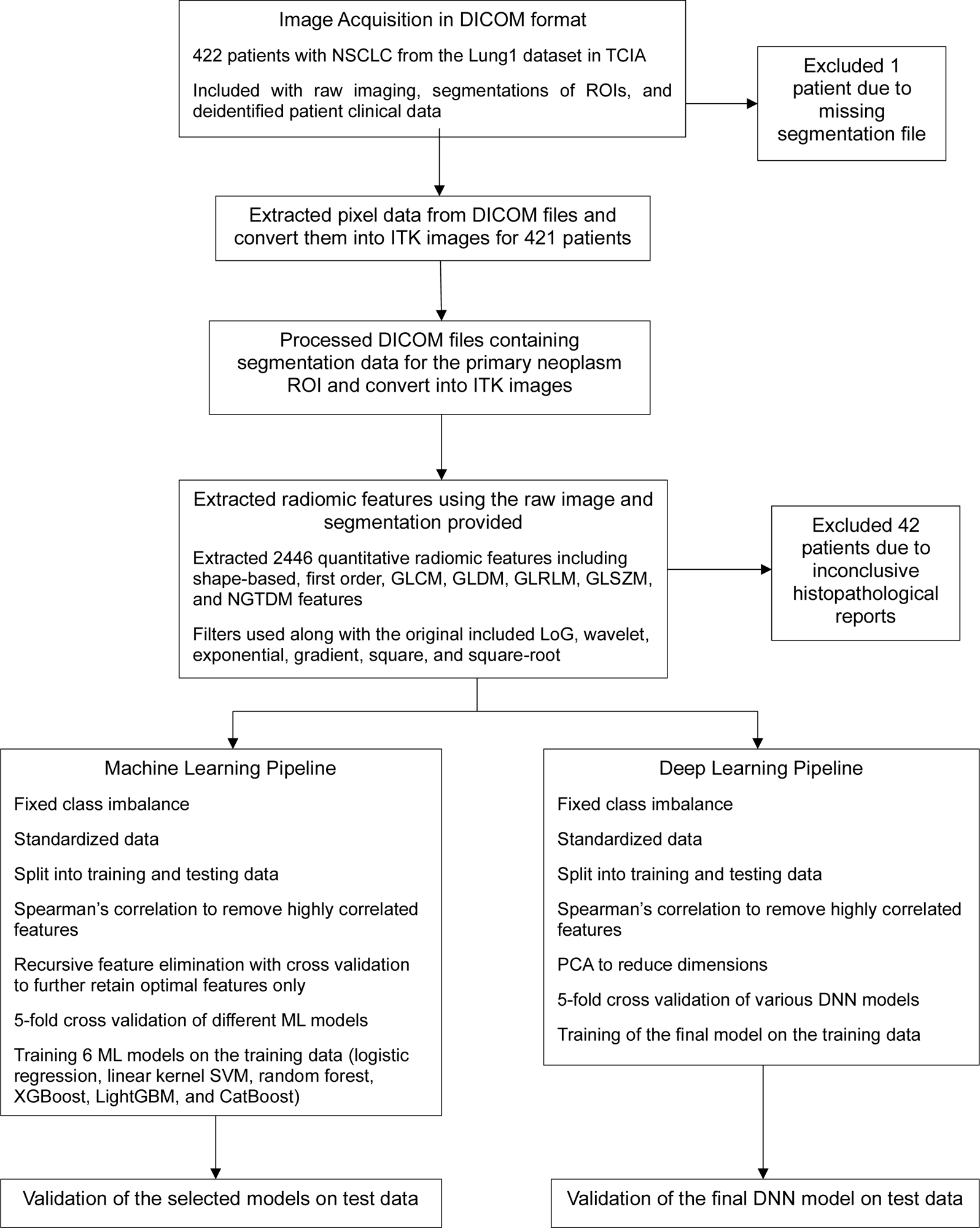
Workflow of radiomic feature extraction and model training

### Data

Data were collected from the NSCLC-Radiomics data collection hosted on The Cancer Imaging Archive.^19^ Care was taken to ensure that there was no overlap between the training and test datasets by splitting data before preprocessing. Data were randomly divided into 80% training and 20% testing sets for machine learning models and 90% training and 10% testing sets for the deep neural network model. Image acquisition and processing were performed using standard clinical protocols for spiral CT scans (3 mm slice thickness) with or without contrast. Fully manual delineation of segments was done by expert radiation oncologists using a standard clinical delineation protocol on fused PET-CT images. Details are provided in original article by Aerts et al.^18^ Clinical variables measured in these patients included age, gender, clinical TNM staging, patient outcome, and survival time. Histological subtype confirmation through biopsy served as the reference standard.

### Pre-processing

Fixed-bin width discretization with a bin width of 32 was used. Original and filtered images (Laplacian of Gaussian [LoG], wavelet, exponential, gradient, local binary pattern [LBP] in 2D and 3D, square, and square root) were used. Scale was normalized to 1. The list of sigma values used for LoG were [0.5, 1.0, 1.5, 2.0, 2.5, and 3.0]. These settings were used to instantiate the feature extractor using the PyRadiomics library.^20^

### Feature Extraction

The hand-crafted features mentioned above were extracted providing a total of 2446 features. Texture, shape, and intensity were the main feature classes.

### Data Preparation

Segmentation data for 1 patient was missing and was dropped. Out of the remaining 421 patients, there was no missing radiomic data. Details about missing clinical data is provided in Table 1. No imputation methods were employed to handle these missing data. Data were split into training and testing groups in an 80:20 ratio. Random oversampling was used to address class imbalance for the machine learning models, while Adaptive Synthetic (ADASYN) oversampling was used for the DNN model. Features were normalized using min-max normalization. For machine learning models, Spearman’s rank correlation was utilized to remove highly correlated features with coefficients ≥0.8. This reduced the total number of features to 321. Recursive feature elimination with cross-validation (RFE-CV) was used to further reduce the number of features to 179 for the machine learning pipeline. For the DNN mode, principal component analysis (PCA) to retain 95% variance was used after Spearman’s rank correlation for dimensionality reduction. This reduced the total number of features to 55 for the deep learning pipeline.

**Table 1:**
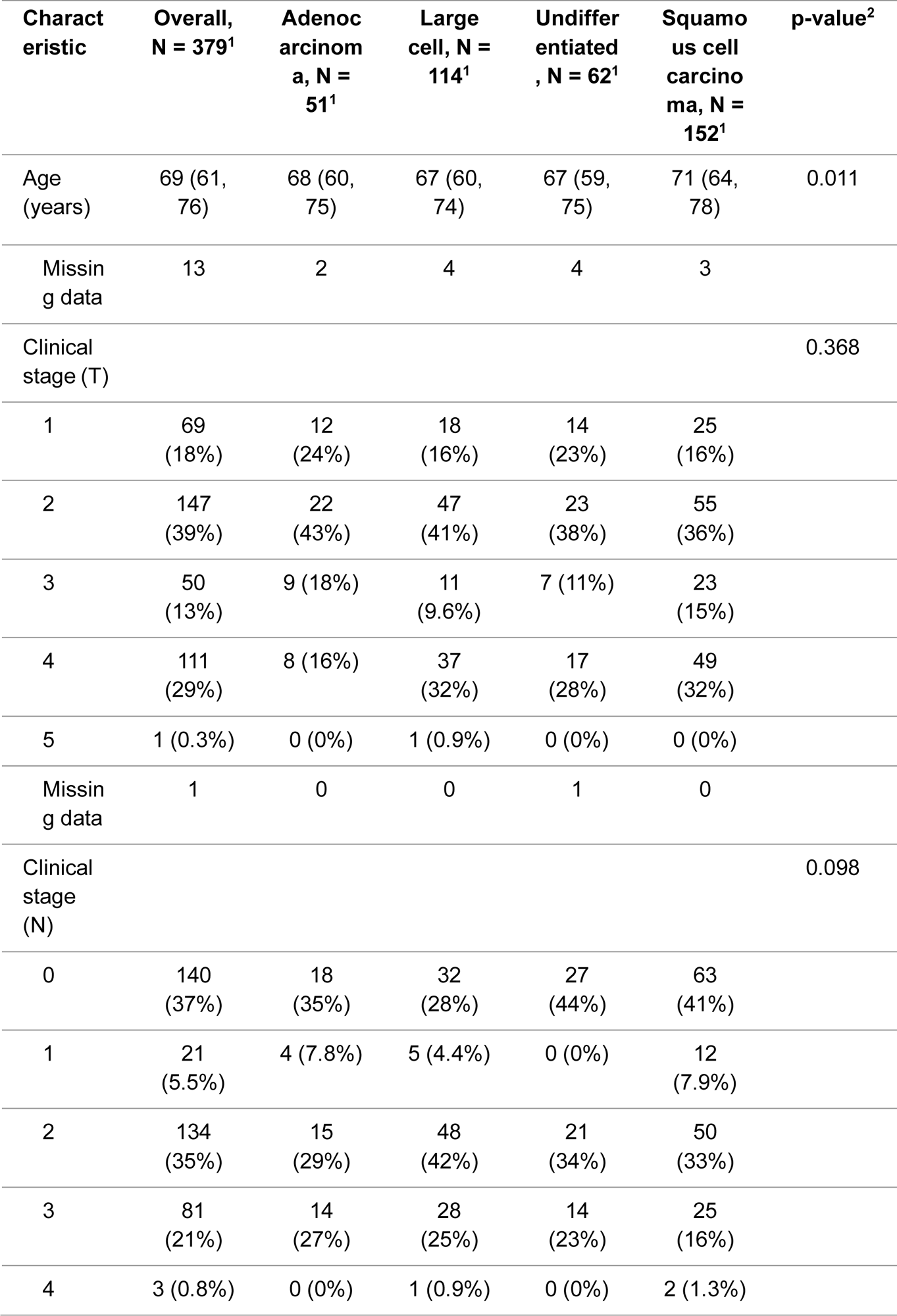
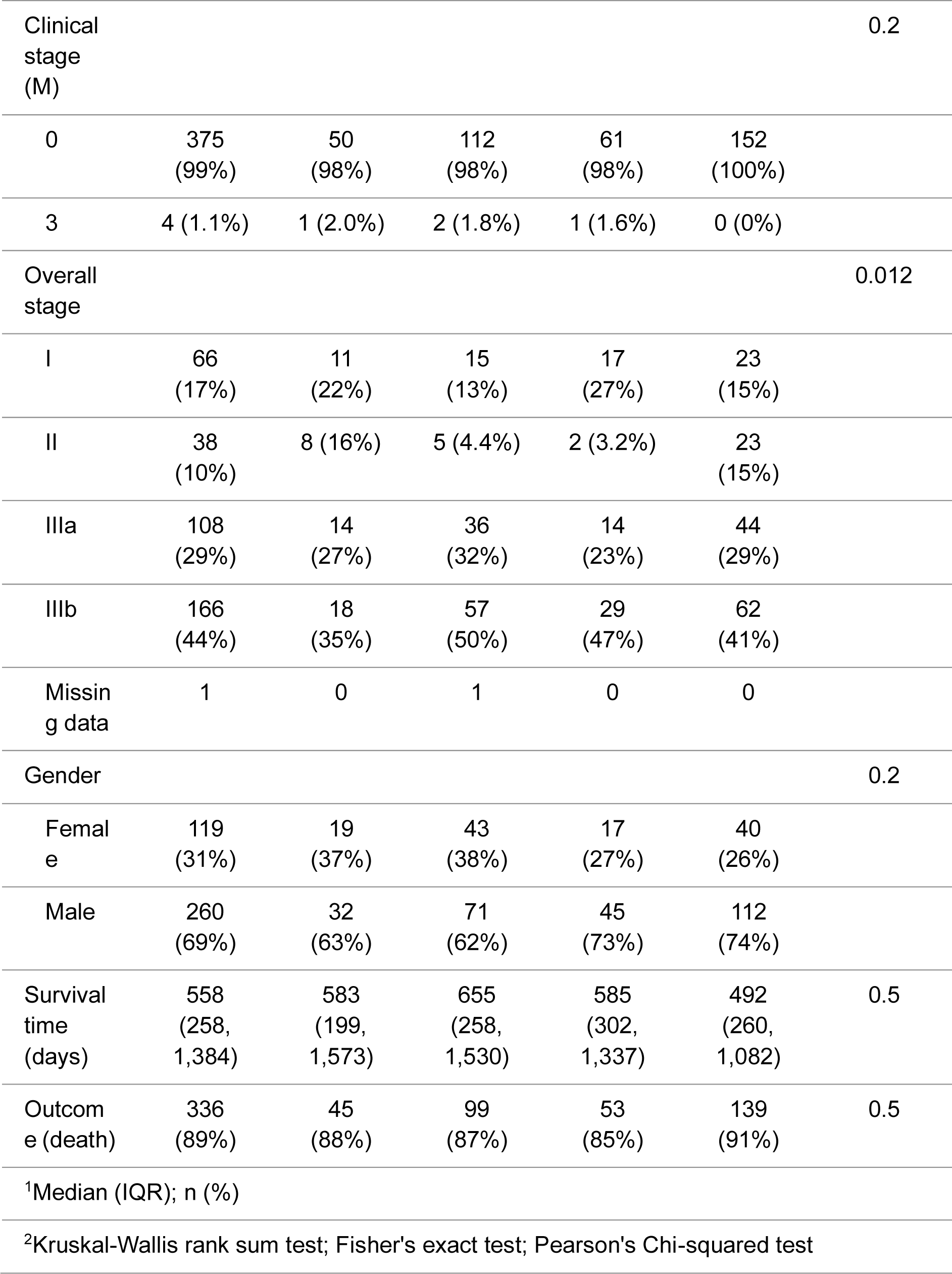
Details about patient and tumor characteristics.

### Machine Learning Modeling

From the scikit-learn library, we decided to use logistic regression (one-vs-rest), support vector machine (SVM) with linear kernel, random forest, extreme gradient boosting (XGBoost), light gradient boosting (LightGBM), and categorical boosting (CatBoost) classification models to predict a multiclass outcome (histology).^21^ We performed 5-fold cross-validation to assess average performance of the models. After modelling the data, we tested their performance on test data.

### Deep Neural Network Modeling

A deep neural network (DNN) was constructed using TensorFlow’s Keras API.^22^ The architecture consisted of five dense layers with decreasing neuron counts from 1024 to 64. Each layer employed L2 regularization and was followed by a LeakyReLU activation function, batch normalization, and dropout layers. The final layer utilized a softmax activation function for classification across multiple categories. The Adam optimizer was employed with an exponential decay learning rate scheduler. We trained the model using categorical cross-entropy loss, monitored for validation loss, and optimized using two callbacks: reduce learning rate on plateau and early stopping. Validation set predictions were subsequently generated.

### Evaluation

Accuracy, precision/positive predictive value (PPV), negative predictive value (NPV), recall (sensitivity), specificity, AUC-ROC were used as performance metrics based on their relevance to classification problems. Confidence intervals were calculated using bootstrapping with 2000 replicates for the machine learning models and 100 replicates for the DNN model.

The methodology was meticulously designed to ensure robustness, repeatability, and transparency in the evaluation of the proposed radiomic analysis technique for predicting NSCLC subtypes. All code and supplementary materials are available upon request.

## Results

In a cohort of 379 lung cancer patients, we analyzed demographic and clinical characteristics based on histological subtypes: Adenocarcinoma (N = 51), Large cell (N = 114), Undifferentiated (N = 62), and Squamous cell carcinoma (N = 152). The median age across the cohort was 69 years (IQR: 61-76), with Squamous cell carcinoma patients being the oldest subgroup with a median age of 71 (IQR: 64-78; p=0.011). Regarding the clinical staging, T-stage distribution showed the majority at stage 2 (39%) and 4 (29%) with no significant variation between histological types (p=0.368). N-stage indicated a high number at stages 0 (37%) and 2 (35%), with a p-value of 0.098. Almost all patients were at M-stage 0 (99%, p=0.2). When analyzing overall cancer stage, the highest proportions were observed in stages IIIb (44%) and IIIa (29%), revealing significant differences between subtypes (p=0.012). Gender distribution highlighted a male predominance (69% male vs. 31% female) across all subtypes, but with no significant variation (p=0.2). The median survival time for the entire cohort was 558 days (IQR: 258-1,384; p=0.5). A majority (89%) of the cases resulted in death, uniformly distributed among the subtypes (p=0.5). (Table 1)

Performances of various machine learning models in predicting the histological subtype of NSCLC were evaluated. Ensemble methods like Random Forest, XGBoost, LightGBM, and Deep Neural Network models showcased the highest accuracies and AUC-ROC values, indicating that they had superior predictive abilities for the histological subtype of NSCLC. (Table 2, Figure 2)

**Figure 2:**
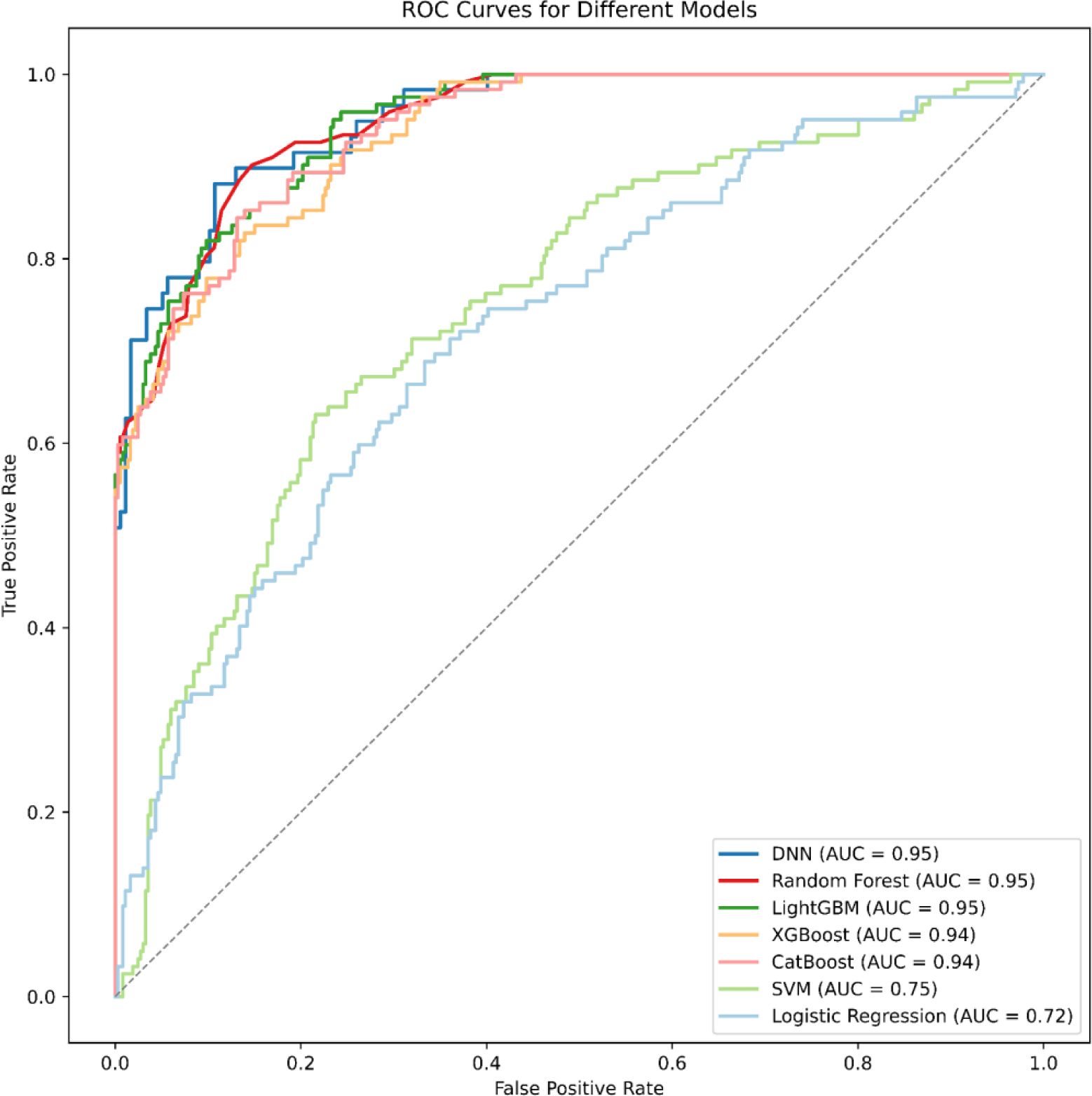
Micro-averaged ROC curves for different models providing performance to determine histological subtypes of NSCLC

**Table 2:**
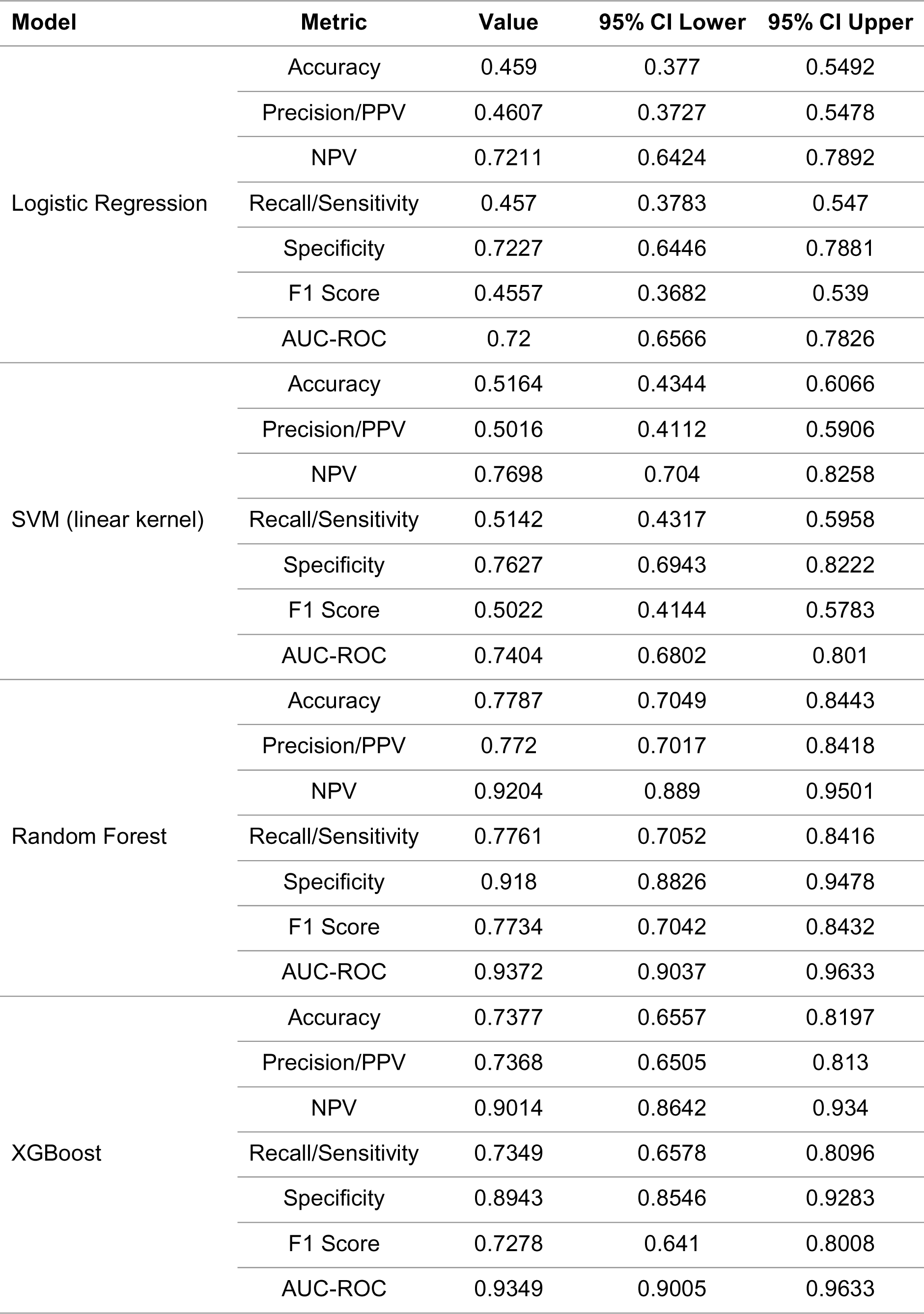
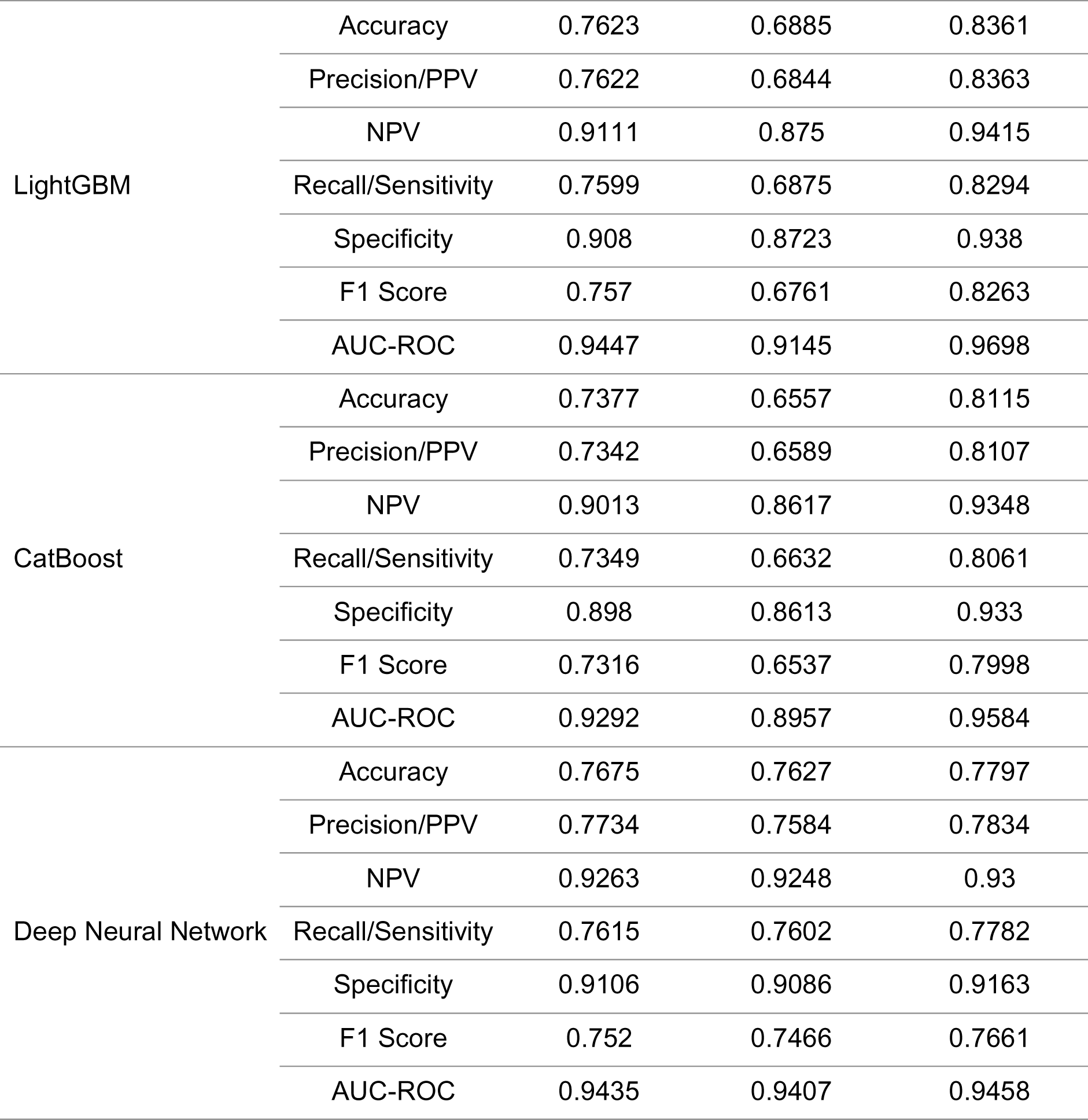
Overall model performance to classify histological subtypes of non-small cell lung cancer.

In one-versus-rest analyses for a multiclass outcome (histology), the performance metrics of different machine and deep learning models on various cancer histological subtypes are provided in Table 3 and 4, illustrated in Figure 3. Almost all models were able to detect adenocarcinoma and undifferentiated carcinoma with high metrics:

**Figure 3:**
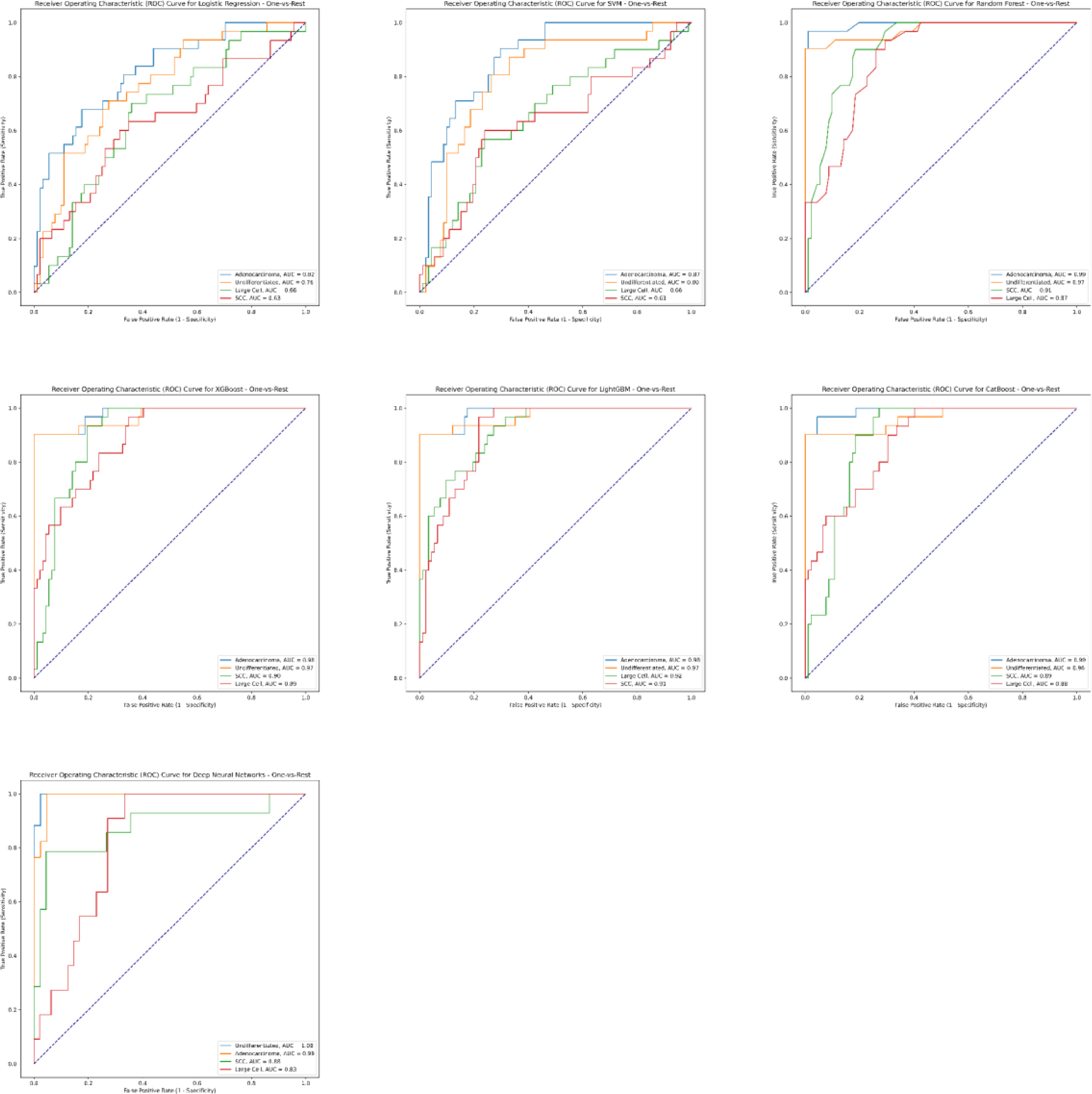
Machine and deep learning model performance to classify non-small cell lung cancers

**Table 3:** One-vs-rest model performance for each histological subtype for the random forest model.

| Cancer Type | Metric | Value | 95% CI Lower | 95% CI Upper |
| --- | --- | --- | --- | --- |
| Adenocarcinoma | Accuracy | 0.9836 | 0.959 | 1 |
|  | Precision/PPV | 0.9677 | 0.8919 | 1 |
|  | NPV | 0.989 | 0.989 | 0.989 |
|  | Recall/Sensitivity | 0.9677 | 0.8929 | 1 |
|  | Specificity | 0.989 | 0.989 | 0.989 |
|  | F1 Score | 0.9677 | 0.913 | 1 |
|  | AUC-ROC | 0.9936 | 0.9786 | 1 |
| Large Cell | Accuracy | 0.8033 | 0.7295 | 0.8689 |
|  | Precision/PPV | 0.6071 | 0.4194 | 0.7813 |
|  | NPV | 0.8617 | 0.8617 | 0.8617 |
|  | Recall/Sensitivity | 0.5667 | 0.3913 | 0.7419 |
|  | Specificity | 0.8804 | 0.8804 | 0.8804 |
|  | F1 Score | 0.5862 | 0.4151 | 0.7213 |
|  | AUC-ROC | 0.8714 | 0.8043 | 0.9254 |
| Undifferentiated | Accuracy | 0.9262 | 0.877 | 0.9672 |
|  | Precision/PPV | 0.8235 | 0.6957 | 0.9394 |
|  | NPV | 0.9659 | 0.9659 | 0.9659 |
|  | Recall/Sensitivity | 0.9032 | 0.7878 | 1 |
|  | Specificity | 0.9341 | 0.9341 | 0.9341 |
|  | F1 Score | 0.8615 | 0.7576 | 0.9444 |
|  | AUC-ROC | 0.9729 | 0.9349 | 1 |
| Squamous Cell | Accuracy | 0.8443 | 0.7787 | 0.9098 |
|  | Precision/PPV | 0.6897 | 0.5172 | 0.8529 |
|  | NPV | 0.8925 | 0.8925 | 0.8925 |
|  | Recall/Sensitivity | 0.6667 | 0.4872 | 0.8294 |
|  | Specificity | 0.9022 | 0.9022 | 0.9022 |
|  | F1 Score | 0.678 | 0.52 | 0.806 |
|  | AUC-ROC | 0.9109 | 0.8598 | 0.957 |

**Table 4:** One-vs-rest model performance for each histological subtype for the deep neural network model.

| Cancer Type | Metric | Value | 95% CI Lower | 95% CI Upper |
| --- | --- | --- | --- | --- |
| Adenocarcinoma | Accuracy | 0.9332 | 0.9311 | 0.9557 |
|  | Precision/PPV | 0.8207 | 0.8152 | 0.8538 |
|  | NPV | 0.9932 | 0.9889 | 1 |
|  | Recall/Sensitivity | 0.9902 | 0.9834 | 1 |
|  | Specificity | 0.9139 | 0.9008 | 0.9328 |
|  | F1 Score | 0.921 | 0.8921 | 0.9293 |
|  | AUC-ROC | 0.9998 | 0.9891 | 1 |
| Large Cell | Accuracy | 0.7936 | 0.7821 | 0.8012 |
|  | Precision/PPV | 0.4327 | 0.421 | 0.4523 |
|  | NPV | 0.8724 | 0.8735 | 0.8819 |
|  | Recall/Sensitivity | 0.4512 | 0.4128 | 0.4514 |
|  | Specificity | 0.8621 | 0.8527 | 0.8823 |
|  | F1 Score | 0.4425 | 0.4289 | 0.452 |
|  | AUC-ROC | 0.8604 | 0.8632 | 0.8735 |
| Undifferentiated | Accuracy | 0.9745 | 0.9741 | 0.9832 |
|  | Precision/PPV | 0.9036 | 0.8923 | 0.9428 |
|  | NPV | 0.9935 | 0.9851 | 1 |
|  | Recall/Sensitivity | 0.9914 | 0.9847 | 1 |
|  | Specificity | 0.9528 | 0.9519 | 0.9834 |
|  | F1 Score | 0.9521 | 0.9428 | 0.9723 |
|  | AUC-ROC | 0.9928 | 0.9812 | 1 |
| Squamous Cell | Accuracy | 0.8523 | 0.8304 | 0.8628 |
|  | Precision/PPV | 0.8542 | 0.7521 | 0.8824 |
|  | NPV | 0.8519 | 0.8423 | 0.862 |
|  | Recall/Sensitivity | 0.4632 | 0.4314 | 0.5017 |
|  | Specificity | 0.9746 | 0.9632 | 0.9823 |
|  | F1 Score | 0.5923 | 0.5538 | 0.6429 |
|  | AUC-ROC | 0.9147 | 0.8924 | 0.9225 |

### 1. Performance on Adenocarcinoma

Random Forest and LightGBM show the highest accuracy with values of 0.9836 and 0.959 respectively. Random Forest achieved the highest AUC-ROC of 0.9936. Both Random Forest and LightGBM have strong precision and recall values, suggesting they can identify Adenocarcinomas effectively.

### 2. Performance on Large Cell

Random Forest provided the best accuracy (0.8033) and AUC-ROC (0.8714). Despite its relatively low recall, the XGBoost model achieves a balanced performance with good precision and F1 Score.

### 3. Performance on Undifferentiated

Random Forest and LightGBM outperform other models in accuracy with 0.9262 and 0.8852 respectively. Random Forest has an excellent AUC-ROC of 0.9729. Both models show good precision and recall, indicating a balanced performance.

### 4. Performance on Squamous Cell

Random Forest achieves the best accuracy (0.8443) and AUC-ROC (0.9109). LightGBM and XGBoost follow closely in terms of performance metrics.

Table 5 provides information on the performance of the random forest model across three different datasets: the original dataset (with class imbalance), an undersampled dataset (with each class having 51 samples), and an oversampled dataset (with each class having 152 samples). Most of the metrics showed lower values when there was undersampling. However, invariably, all the metrics showed improvement after oversampling and led to the best performance of the classifier. The model was validated on the test set which was otherwise unexposed to preprocessing and model training.

**Table 5:**
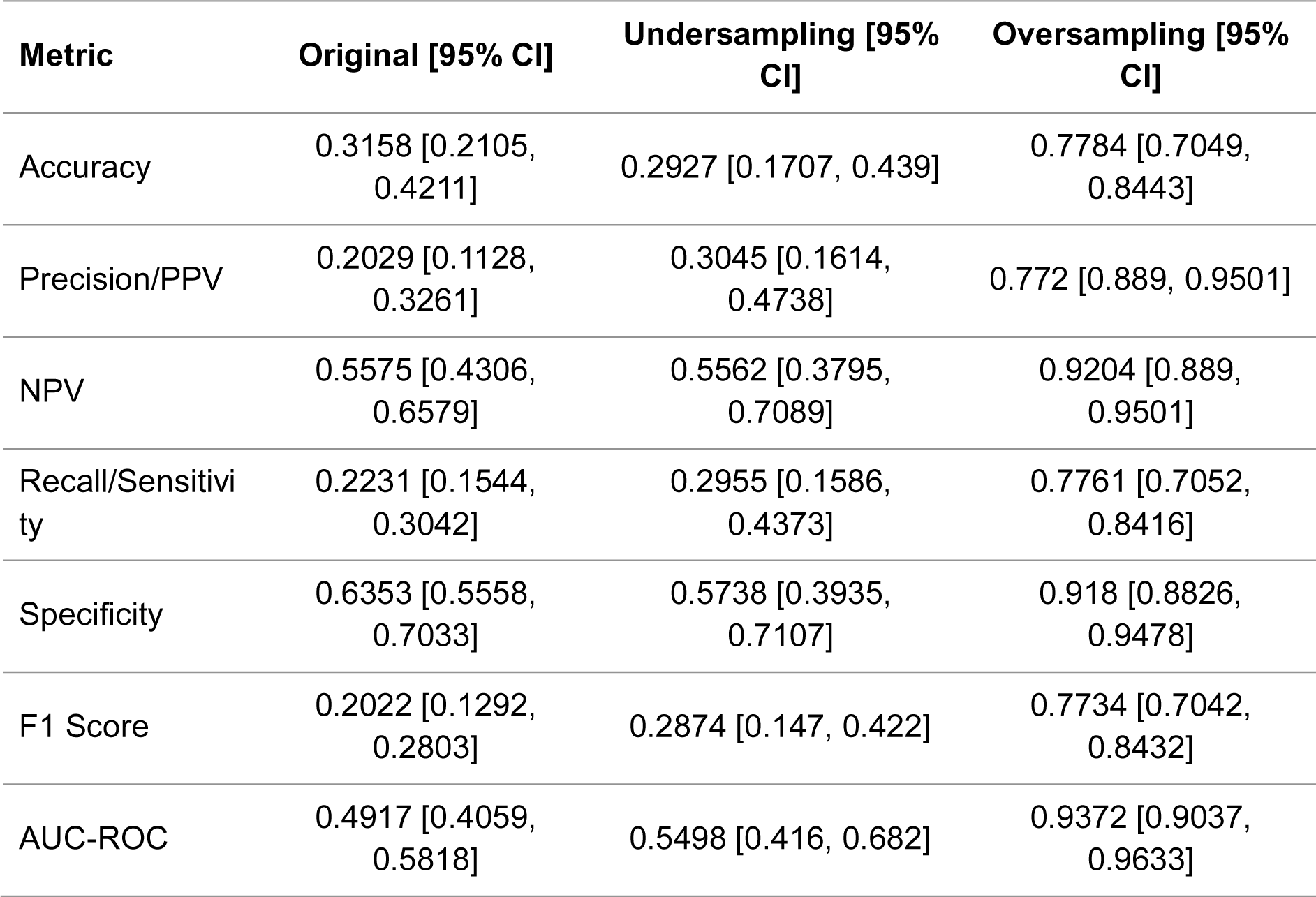
Overall random forest model performances for 3 pipelines – original sampling (unbalanced histological classes), random under sampling (balanced histological classes), or random over sampling (balanced histological classes)

### Discussion

Lung cancer remains a major global health challenge, with Non-Small Cell Lung Cancer (NSCLC) forming a significant proportion of cases.^2^ Our study was a technical task-specific evaluation focused on building and evaluating the predictive capabilities of machine and deep learning models to predict the histological subtype of NSCLC from CT scans using hand-crafted quantitative radiomic analysis. We successfully demonstrated the potential of quantitative radiomic analysis in tandem with machine and deep learning techniques to provide accurate, non-invasive prediction of NSCLC histological subtypes using CT images. Specifically, our findings indicated that ensemble methods such as Random Forest and LightGBM as well as deep learning using DNN outperformed other models, particularly in the detection of adenocarcinoma and undifferentiated carcinoma.

This study offers a fresh perspective on combining radiomic approaches with artificial intelligence (AI) techniques for prediction modelling. Our results consolidate findings supporting the transformation of diagnostic processes for complex classifications. The history of radiomics has undergone substantial growth, with only a few features being extracted and used initially to usage of advanced computational techniques to extract upwards of 2000 features.^18,23^ The use of radiomics has been consistently showing tremendous application in the diagnosis of malignancies, such as lung cancer.^24^ Previous studies have documented that with similar samples sizes such as ours, the AUCs obtained ranged from 0.71 to 0.87 to detect histologic subtype of NSCLC.^25^

Historically, the standard technique for NSCLC subtype identification necessitated an invasive biopsy procedure followed by a meticulous histopathological examination. Such methods, which are gold-standard currently, pose potential risks and can be time-consuming.^26^ Recent advancements in the field of radiomics and artificial intelligence have hinted at the possibility of a more refined, non-invasive, and time-efficient approach.^27^ A study by Wu et al. studying wavelet-based features achieved a AUC-ROC of 0.72 using Naïve Baye’s classifier.^28^ Ferreira et al. achieved an AUC-ROC of 0.92 in detecting histology of lung cancer.^29^ Radiomic analysis also proved to have a high AUC-ROC in detecting epithelial ovarian carcinoma subtypes on CT with AUCs or 0.836.^30^ In contrast to most of such studies, our study has achieved AUC-ROCs of 0.95 in detecting the histological subtype of NSCLC. Moreover, these studies have extracted variable numbers of features ranging from 107 to 1160 radiomic features, using 3D-Slicer and MATLAB to do so.^31,32^

The distinct edge our study offers over previous research lies in the using a standard PyRadiomics pipeline to extract all features possible using different filters to offer precise subtype predictions based purely on CT scans. Further, we have applied a filter-based feature selection method followed by a wrapper-based method to ensure retaining only the most useful features which contribute to the model. Lastly, 5-fold cross validation of each of our models ensured best and most optimal selection.

The implications of this study on clinical practice are multifold:

#### Non-invasive Diagnostic Tool

With our proposed methodology, there arises a potential to significantly minimize or even negate the need for invasive biopsies in certain cases, thereby reducing associated procedural risks and patient discomfort.

#### Efficiency and Accuracy

Our method ensures quicker and accurate subtype predictions based on initial CT scans. This can potentially shorten the diagnostic journey, leading to faster, targeted treatments and better patient outcomes.

#### Standardized Diagnostics

Harnessing the quantitative nature of radiomics ensures a more objective diagnostic criterion, effectively reducing interobserver variability which often plagues traditional methods.

#### Optimized Resource Allocation

In a busy clinical setting, relying on automated tools like the ones we propose can help reduce human error, optimize resource utilization, and manage patient flow more effectively.

While our findings are promising, they are not without limitations. The sample size was relatively small with 422 patients in the dataset. Furthermore, 43 patients did not have conclusive histopathological diagnoses for their NSCLC. Class imbalance necessitated balancing methods including random oversampling and ADASYN. Although these are robust methods, these are randomly generated and synthetically generated samples respectively. Finally, different feature selection methods were employed in the machine learning and deep learning models to ensure best model performances after trial and error.

The compelling results from this study further unravel the potential of radiomic analysis in the classification of cancer, specifically NSCLC, subtypes. While the current study offers promising insights, there are several avenues to further enhance and solidify these findings in the future:

#### Larger Datasets

In order to improve the robustness of the models, further studies should aim to incorporate larger and more diverse datasets. Currently, we observed that we were able to get optimal results after oversampling. This might suggest that the current sample size was insufficient to train the model without oversampling to fix class imbalance. This will ensure that the models are better generalized and can accurately detect NSCLC subtypes across various populations and clinical scenarios.

#### Real-time Application

Exploring the feasibility of integrating these machine learning models into real-time diagnostic platforms can revolutionize clinical decision-making processes. Such integration will allow radiologists and oncologists to make instant, evidence-based decisions regarding patient management.

#### Expanding Modalities

Beyond CT scans, future studies could explore the potential of combining radiomic data from other imaging modalities such as MRI and PET scans. This multi-modal approach might capture a more comprehensive picture of tumor characteristics, leading to even more accurate classification.

#### Temporal Analysis

Evaluating how radiomic features evolve over time could provide insights into tumor progression and its correlation with histological transformation, if any. This could further aid in predicting tumor behavior and response to treatment.

#### Personalized Treatment

With accurate histological subtype classification, future research could delve deeper into tailoring specific therapeutic regimens based on the identified subtype, leading to personalized treatment plans and potentially better patient outcomes.

#### Integration with Genomics

By merging radiomic data with genomic information, there is potential to uncover relationships between imaging features and molecular signatures. This could pave the way for more comprehensive diagnostic tools that consider both the physical and molecular landscape of the tumor. Furthermore, it might be interesting to study whether the type of genetic mutation in pro-oncogenes/tumor-suppressor genes is associated with distinct radiomic signatures.

#### Model Interpretability

As the field of machine learning grows, there’s increasing emphasis on model interpretability. Future research should focus on developing models that not only predict accurately but also provide insights into which radiomic features are most indicative of specific histological subtypes.

In conclusion, our study attempts to instantiate a radiomic analysis pipeline to extract standard features from lung CT scans and run ensemble machine learning classifiers and deep learning models to predict histology of the lesion. As the age of precision medicine advances, such innovations aim to redefine how we approach, diagnose, and eventually treat diseases as multifaceted as NSCLC.

## Open Science

Images and segmentation data are available from NSCLC-Radiomics in The Cancer Imaging Archive.^19^

Radiomic feature data extracted by us, pre-processing scripts and settings, source code for modeling, and final model files will be shared by us provided a reasonable request is made to the corresponding author.

A ready-to-use system in the form of a web-based application where users can upload DICOM files and segmentation files is being developed. Kindly contact the corresponding author to know more about the status.

## Data Availability

All data produced are available online at https://www.cancerimagingarchive.net/collection/nsclc-radiomics/

## Acknowledgements

This project was supported by the Institute of Electrical and Electronics (IEEE), Engineering in Medicine and Biology Society (EMBS), Indian Institute of Technology, Kharagpur.

